# Estimating excess mortality during heat waves using a causal inference framework and non-randomized time series

**DOI:** 10.64898/2026.10.13.26365467

**Authors:** Yasmine Mabene, Lauren Mock, Luke Miratrix, Young Lee, Marie-Abèle Bind

**Author notes:** **Correspondence** Yasmine Mabene, Department of Computer Science, Stanford University, Stanford, CA, 94305, USA. Equally contributing authors (Yasmine Mabene, Lauren Mock). Equally contributing authors (Luke Miratrix, Young Lee, Marie-Abèle Bind). **Funding information** Office of the Director, National Institutes of Health, Grant/Award Number: DP5OD021412. The content is solely the responsibility of the authors and does not necessarily represent the official views of the National Institutes of Health.

## Abstract

Researchers must often rely on observational data to draw conclusions about the causal relationship between an exposure and an outcome. This is common in environmental epidemiology research, where the assignment to an exposure can be unethical or impossible. Consider a setting in which we seek to estimate excess mortality during heat waves, but are unable to randomize individuals to moderate or extreme heat. We propose to follow a causal framework for non-randomized time series that uses what we call *historical time series matching* to construct hypothetical randomized experiments, in which background covariates are balanced. We applied this framework and assessed the following causal question: Are daily mortality counts among elderly Americans higher during hot versus warm summer time periods? We addressed this causal question by drawing frequentist, Fisherian, and Bayesian inferences for five American cities. We estimated the average treatment effect of heat on three-day mortality with 95% Fisherian intervals to be 0.5 [-3.6, 4.4] deaths in Pittsburgh, 3.7 [0.7, 6.6] deaths in Seattle, 12.5 [6.1, 18.9] deaths in Chicago, 8.5 [1.4, 15.6] deaths in Los Angeles, and 20.3 [11.2, 29.5] deaths in New York.

## 1 INTRODUCTION

Global climate change is a substantial threat to human health ^1^. In the United States, the average temperature has increased between 1.3 and 1.9°F since record-keeping began in 1895, with a total projected increase of 3°F to 10°F by the end of this century ^2^. In addition to increases in average temperature, extreme heat events have become increasingly frequent and intense. The Intergovernmental Panel on Climate Change attributes increases in heat-related mortality to these heat events ^3^, which are the leading cause of weather-related mortality in the United States ^4^. Extreme heat can interfere with the body’s temperature regulation, resulting in the exacerbation of chronic conditions and the onset of several new health complications, leading to morbidity and mortality ^2^. Although extreme heat events are becoming more widespread, the human health impacts of rising temperatures are not evenly distributed. Extreme heat events disproportionately affect the elderly population and marginalized groups^1,2,5,6^. Those living in urban areas are especially vulnerable due to the urban heat island effect, which explains why dense cities are hotter than surrounding rural areas ^7,8^. Additionally, the synergistic health effects of high temperatures and air pollution further contribute to elevated risks in these areas ^4^.

Previous epidemiological studies have identified a J-, V-, or U-shaped association between temperature and mortality ^9,10^. Several studies have linked rising temperatures with increased risks for natural, cardiovascular, and respiratory mortality ^5,11,12^, and others indicate that daily mortality counts increase during heat waves ^13,14^.

Determining the temperature-mortality relationship from medical records may underestimate the number of temperature related deaths due to lack of consistent diagnoses ^2^. More commonly, temperature-mortality studies utilize time series and case-crossover methods ^15^. The time series approach involves comparing mortality outcomes at different exposure levels for a set of intervals over a period of time. These studies frequently incorporate distributed lag nonlinear models and generalized additive models to investigate the relationship between temperature and mortality ^16–18^. Typically, smoothing functions such as natural splines are used to control for potential confounding factors including seasonality and humidity. However, existing correlations between nonlinear functions or small regression coefficients can lead to biased estimates in these models ^19^. Additionally, *a posteriori* selection for the parametric form of the functions in these models inflates type I errors and results in undercoverage in confidence intervals ^20–23^.

The case-crossover design resembles a case-control study, but cases serve as their own reference, allowing for the control of time-invariant, individual-level confounding factors. When examining temperature-mortality relationships, case-crossover studies often rely on estimated coefficients from conditional logistic regression models to estimate causal effects ^24,25^. However, these models can produce misleading findings as a result of selection bias and overlap bias ^24,26–28^. Estimates based on maximum likelihood estimation may also be heavily biased for studies with small sample sizes or with high or low prevalence risk factors ^24^.

To estimate the causal effect of hot temperatures on mortality, we rely on the Neyman-Rubin Causal Model framework, in which we depart from traditional association modeling, and instead design the observational study prior to its analysis ^29,30^. We obtain daily measurements of mortality, meteorological variables, and air pollution concentrations in five major cities in the United States. We conceptualize a plausible, hypothetical randomized experiment in which three-day time periods are randomly assigned to be exposed or unexposed to extreme heat. We then identify comparable exposed and unexposed time periods with respect to certain covariates selected *a priori*, implement a maximum bipartite matching algorithm to identify the best 1:1 pairs of exposed and unexposed time periods, and conduct our analysis on these matched pairs ^31–34^. By following this framework, we follow a preferred method for determining causal effects ^35^, limiting the bias that occurs due to confounding in traditional time series approaches^36^. Additionally, the randomization-based inference approach does not depend on asymptotic distributional assumptions ^37,38^.

After designing this experiment, we assess the temperature-mortality relationship with frequentist, Fisherian, and Bayesian approaches. Our study provides estimates and uncertainty intervals for the causal effect of extreme temperatures on mortality from each approach. Furthermore, the causal inference framework implemented here can be utilized in other observational studies to estimate the effects of environmental exposures on health outcomes^32,33,39^.

## 2 METHODS: A CAUSAL INFERENCE FRAMEWORK

We obtained daily counts of mortality, maximum and minimum temperatures, dew point temperature, relative humidity, and air pollution concentrations (i.e., O_3_, NO_2_, CO, SO_2_, PM_10_, and PM_2.5_) between 1987 and 2000 in five American cities: Pittsburgh, Seattle, Chicago, Los Angeles, and New York. These data were collected through the National Morbidity, Mortality, and Air Pollution Study (NMMAPS). Total daily mortality counts in the NMMAPS dataset are grouped into three age categories: under age 65, between ages 65 and 74, and 75 years or older. As the elderly are much more susceptible to extreme heat events than younger people ^6^, the study population was limited to those aged 65 and older. The number of deaths due to certain specific causes was available but not incorporated into this study, which focuses on all-cause mortality. Other characteristics of those who died each day were unavailable. The study’s time period was limited to days in June, July, August, and September; all days during these months will be referred to as “summer” days. Of the 8,540 summer days between 1987 and 2000 across five cities, the total mortality count among those 65 and older was unavailable for 11 days, and the maximum temperature was unavailable for one day. These days were removed from the analysis. We followed a causal inference framework ^30,33,34,38,39^, adapted for observational time series ^32^.

### 2.1 Stage 1: Conceptual Intervention

We seek to assess the following causal question: Are daily mortality counts among elderly Americans higher during hot versus warm summer time periods? Because we cannot conduct an experiment that randomly assigns summer days to be either hot or warm, we instead set up a plausible, *hypothetical* randomized experiment and consider the Neyman-Rubin Causal Model ^29,30,32,39,40^. In this analysis, we estimate effect sizes for each city separately to account for city-specific differences in age structure, weather patterns, air pollution, and cooling infrastructure.

Because heat events are typically defined by multiple consecutive days with higher-than-usual temperatures, we investigate three-day time periods. We refer to each rolling three-day period comprised of Day 1, Day 2, and Day 3 as an “experimental unit.” We refer to the days preceding each experimental unit as lag days, counting backwards; the day before an experimental unit is denoted by Lag 1, two days before the experimental unit by Lag 2, etc.

Consider a period of three consecutive summer days denoted by *i* (where *i* ∈ {1, …, *N*}) in city *c*. For any day *i* in city *c*, we denote the maximum daily temperature by temp.max_*i,c*_ and a dichotomous, city-specific treatment by *W*_*i,c*_. We define a “treated” or hot time period in a given city *c* to be three consecutive summer days, for which temp.max_*i,c*_ is above that city’s summer median daily maximum temperature. We define a “control” or warm time period in a given city *c* to be three consecutive summer days, for which temp.max_*i,c*_ is below or equal to that same threshold. Any other three-day period is considered to have an undefined treatment and is excluded from our hypothetical randomized experiment. See Figure 1 for examples of control, treated, and undefined periods.

**FIGURE 1.**
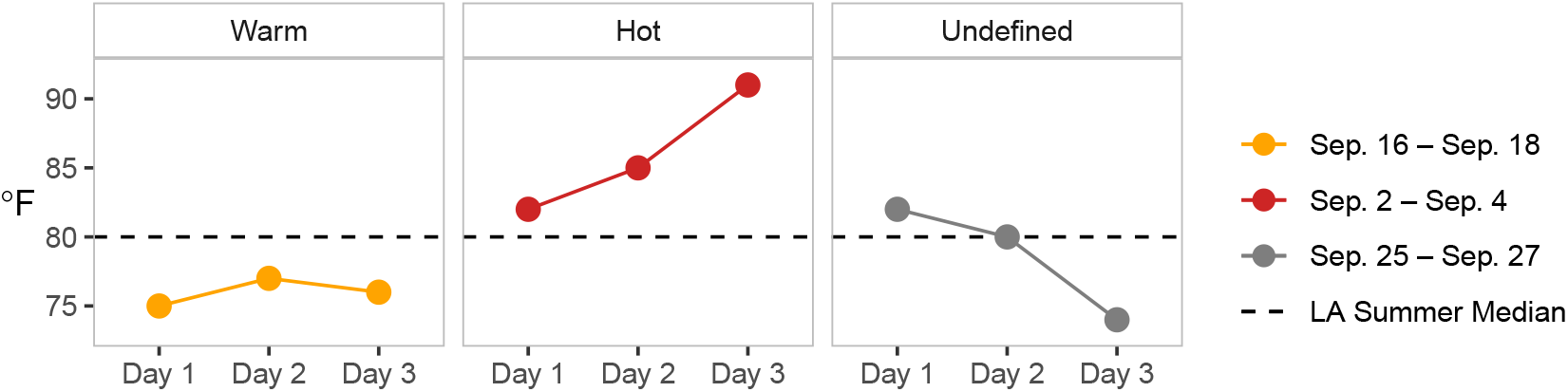
Examples of warm (control), hot (treated), and undefined three-day periods in 1990 in Los Angeles as defined in Equation 1.

Our dichotomous treatment indicator, *W*_*i,c*_, is then defined in the following way for each rolling three-day period *i* in city *c*:

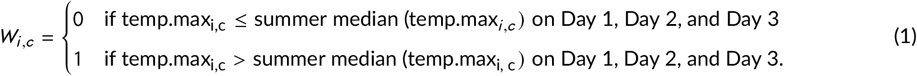

The median daily maximum temperatures (temp_max_) across summer days in each city are reported in Table 2. While Pittsburgh, Chicago, Los Angeles and New York have similar medians around 80°F, Seattle has a notably lower median of 72°F.

*Y*_*i,c*_, our outcome of interest, is the total number of deaths among people aged 65 and older in a given city *c* during a three-day period *i*. If a three-day period is a control unit (warm), then the treatment indicator *W*_*i,c*_ = 0, and the observed number of deaths is *Y*_*i,c*_ (*W*_*i,c*_ = 0). Similarly, if the three-day period is a treated unit (hot), then *W*_*i,c*_ = 1, and the observed number of deaths is *Y*_*i,c*_ (*W*_*i,c*_ = 1).

### 2.2 Stage 2: Design of the observational study

In the next stage of this framework, we seek to obtain a subset of the data, for which we can assume that treatment assignment is unconfounded given the measured covariates, *X*_*i,c*_; i.e., *P* (*W*_*i,c*_ | *Y*_*i,c*_ (*W*_*i,c*_ = 0), *Y*_*i,c*_ (*W*_*i,c*_ = 1), *X*_*i,c*_) = *P* (*W*_*i,c*_ | *X*_*i,c*_) ^41^. In other words, we seek to identify time periods for which the treatment assignment is independent of the joint potential outcomes, conditional on a set of observed confounders ^37^.

Here, we use matching to approximate the unconfoundedness assumption. We identify groups of treated and control units with similar lag days, meaning that the days preceding the treated and control three-day periods are similar with respect to observed potential confounders. For each potential confounder, we set the maximum discrepancy allowed between treated and control periods in order to consider them comparable. These thresholds are listed in Table 5. Stricter matching requirements lead to better covariate balance between treated and control units, but fewer matched pairs to evaluate our causal question.

We then employ a maximum bipartite matching algorithm ^32–34^ to identify the most similar pairs of treated and control units among all those that meet the requirements set in Table 5. Our study population is therefore limited to the pairs of treated and control units that are selected through this matching process ^31–34^. This causal inference step, which we call “historical time series matching,” ensures that the days preceding matched treated and control units are as similar as possible, thus mimicking a randomized experiment. This algorithm accounts for mortality differences by year, season, and day of the week, as well as potential lagged effects of temperature, humidity, and pollutants on mortality. For example, on August 8^th^, 9^th^, and 10^th^ 1987, the daily maximum temperatures in Chicago were each above Chicago’s summer median maximum temperature (80°F), making these three days a “treated” time period. We aim to match this treated period with a corresponding control period—three consecutive summer days that were each below or equal to 80°F in Chicago. This control period must span the same three days of the week (i.e., Saturday, Sunday, and Monday) as the treated period and also fall within three weeks of the treated period. This limits potential matching control periods to July 18^th^—20^th^, July 25^th^—27^th^, and August 1^st^—3^rd^ 1987 in the three weeks preceding the treated period, and August 15^th^—17^th^, August 22^nd^—24^th^, and August 29^th^—31^st^ 1987 in the three weeks following the treated period. In addition, we require NO_2_ and O_3_ concentrations to be within 5 *µg* /*m*^3^ on the single days preceding the three-day treated and control periods (Lag 1). If none of the six potential control periods meet these criteria, the treated period cannot be matched. If a single control period fits these criteria, it is matched with the treated period. If multiple control periods fit these criteria, the maximum bipartite matching algorithm identifies the control period that is most similar to the treated period, and only the control period is matched with the treated period. We then limit our analysis to only those treated and control units that were matched, with equal numbers of treated and control units.

### 2.3 Stage 3: Statistical Analysis

After designing the observational study in this hypothetical randomized experiment, we apply three inferential frameworks to assess our causal question: frequentist (Neyman), randomization-based (Fisher), and Bayesian (Rubin). In each framework, we assume that the stable unit treatment value assumption (SUTVA) ^42^ holds, which means the treatment assignments for the experimental units cannot affect the observed outcomes for any given unit and each experimental unit can only receive its given treatment (i.e., hot vs. warm).

#### 2.3.1 Frequentist inference

##### Neymanian inference

###### Estimands

For the *i*^*th*^ time period in a given city *c*, the unit-level causal treatment effect on the additive scale, *τ*_*i,c*_, is defined as follows:

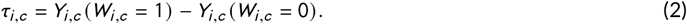

However, we are unable to observe both *Y*_*i,c*_ (*W*_*i,c*_ = 1) and *Y*_*i,c*_ (*W*_*i,c*_ = 0) for a single period; this is the fundamental problem of causal inference ^43^. Recall that if a period is hot, the number of deaths we observe during that period is *Y*_*i,c*_ (*W*_*i,c*_ = 1); if it is warm, we observe *Y*_*i,c*_ (*W*_*i,c*_ = 0). For this reason, we instead estimate the average difference between the number of deaths we would have observed had time period *i* in city *c* been hot (treated) and the number of deaths we would have observed had the same time period *i* in city *c* been just warm (control), i.e., the average treatment effect in each city, which we denote by *τ*_*c*_. *τ*_*c*_ is also the average difference between the potential outcomes *Y*_*i,c*_ (*W*_*i,c*_ = 1) and *Y*_*i,c*_ (*W*_*i,c*_ = 0) across all periods *i* (*i* ∈ {1, …, *N*}) in a city *c*:

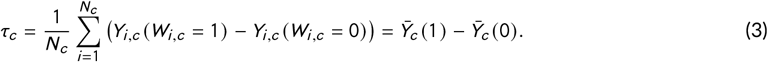

###### Point estimate

The estimated average treatment effect in city *c*, 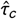, is the difference between the average number of deaths during hot periods and the average number of deaths during warm periods:

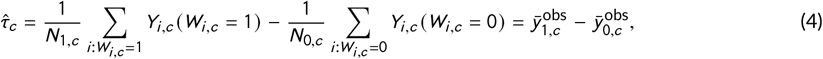

where *N*_1,*c*_ is the number of matched treated periods in city *c* and *N*_0,*c*_ is the number of matched control periods in city *c*. Because we use one-to-one matching, *N*_1,*c*_ = *N*_0,*c*_ = *N*_*c*_ /2, where *N*_*c*_ is the total number of matched periods in city *c*.

For the *k*^th^ pair of treated and control time periods (i.e., *k* ∈ {1, …, *N*_*c*_ /2}), let *d*_*k, c*_ denote the observed cityspecific difference between the number of deaths during the treated time period (denoted by 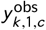) and the control time period (denoted by 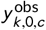):

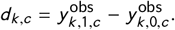

Using the above notations, the estimated average treatment effect 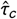 can also be expressed as 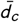, the average of 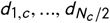

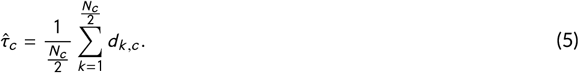

###### Sampling variance estimator

Previous papers have determined the variance properties of 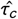^44–46^, allowing us to construct uncertainty intervals for this estimate. Let 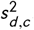, the sample variance of *d*_1,*c*_, …, *d*_*k, c*_ be defined in the following way:

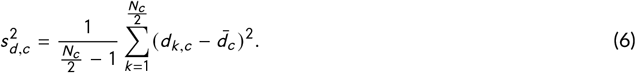

If the treatment effect is the same for each pair *k*, an unbiased estimate of var 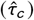 is:

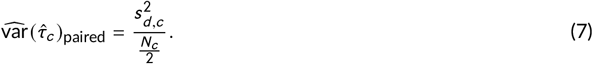

Note that if the treatment effect is not the same for each pair, 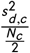 is a conservative estimate of 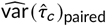. Assuming a constant unit-level treatment effect, we can also estimate the variance without considering pairing:

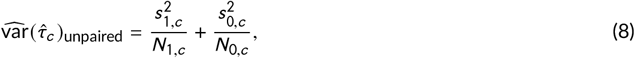

where 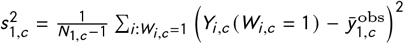 and 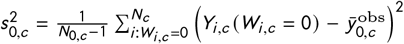

###### Uncertainty interval

We estimate 95% confidence intervals for *τ*_*c*_ as:

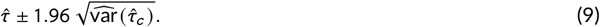

##### Poisson Regression

Historically, there has been a trend toward employing regression analysis to uncover causal effects. During this time, researchers began to shift their focus away from meticulous planning and thoughtful consideration, favoring instead the quicker and more straightforward solutions offered by statistical methods. These methods involved regressing an observed health outcome on an environmental exposure of interest and background covariates (see discussions in ^47^). One alternative frequentist approach is to use Poisson regression, which models the count of deaths over a three-day period. For each city, we fit three separate Poisson regression models:

**Model 1**. A model with only treatment as an explanatory variable:

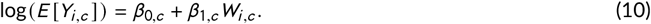

**Model 2**. A model using treatment and pair as explanatory variables:

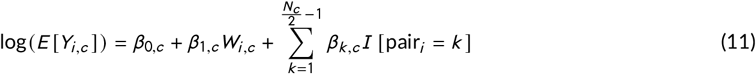

**Model 3**. A model using treatment, pair, month, as well as O_3_ and NO_2_ concentrations on the day preceding the three-day period (Lag 1) as explanatory variables:

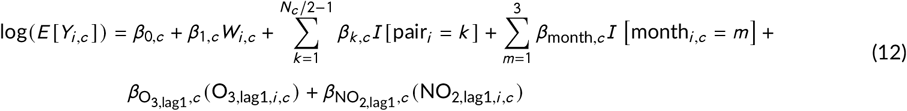

We estimate 95% confid(ence intervals for the risk ratios, 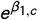, using the estimated Poisson regression coefficients and standard errors exp 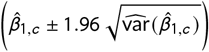.

#### 2.3.2 Fisherian inference

##### Fisher-exact p-value

We also use a Fisherian approach to test city-specific sharp null hypotheses (*H*_0,*c*_) stating that the three-day city-specific mortality is the same during hot and warm time periods. Using this framework, we proceed with a stochastic proof by contradiction of the city-specific sharp null hypotheses ^48,49^, which are defined as the following:

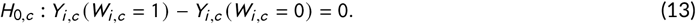

Thus, we assume that the observed and unobserved potential outcomes for each three-day period in a given city *c* are the same and seek a stochastic, empirical contradiction.

As our test statistic, we choose 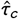, the estimated average treatment effect in city *c* defined in Equation (4) in the Neymanian inference section. We construct the null randomization distribution, i.e., the set of test statistics calculated under *H*_0,*c*_ for all possible treatment allocations in the hypothetical city-specific experiment. Because the experimental units (i.e., the sets of three-day hot and warm days in a given city *c*) are paired using one-to-one matching, we assume a paired-randomization design, such that there are 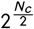 possible treatment allocations, where *N*_*c*_ /2 is the number of matched pairs for each city. Running all possible treatment allocations is quite computationally intensive, thus we approximate the null randomization distribution using 100,000 treatment allocations.

Using the null randomization distribution, we calculate the one-sided Fisher-exact p-value, which is defined as the following:

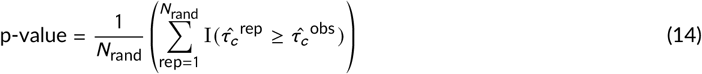

where *N*_rand_ is the total number of calculated test statistics under *H*_0,*c*_ (here, 100, 000), 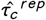 is the estimated average treatment effect in city *c* after randomization, and I is the indicator function (i.e., I(*x*) = 1 if *x* is true, 0 otherwise). The Fisher-exact p-value is the proportion of test statistics as or more extreme than the observed test statistic, 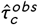.

##### Fisherian intervals

To determine the range of effect sizes consistent with our data, we compute Fisherian intervals (FIs) for a common additive treatment effect ^38^. Unlike (Neymanian) confidence intervals, which provide estimated uncertainty of average treatment effects, Fisherian intervals consist of a range of city-specific *unit*-level treatment effects *τ*_*i,c*_ that are assumed constant ^32^. Similar to Fisher-exact p-values, Fisherian intervals do not rely on asymptotic assumptions.

###### Additive scale

Fisher intervals consist of a range of effect sizes, *τ*_diff_, for which the associated sharp hypothesis, stating that the unit-level causal effect is equal to *τ*_diff_, is not rejected. To obtain Fisherian intervals, we generate null randomization distributions and calculate Fisher-exact p-values for a set of sharp hypotheses contrasting the unit-level potential outcomes on the additive scale ^41^. We use the following hypothesis to calculate Fisherian intervals for *τ*_*i,c*_ defined in Equation 2:

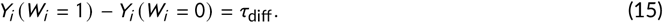

###### Multiplicative scale

Additionally, we use the above hypothesis to estimate Fisherian intervals for the unit-level risk ratios. We use the exponent of the estimated coefficient from the Poisson regression model, i.e., 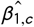 from model 1 (Equation 10), as our test statistic. Using *α* = 0.05 as our significance level, we retrieve two effect sizes such that the Fisher-exact p-values of their respective null randomization distributions equal 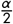 and 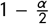. These two effect sizes form the bounds of the Fisherian interval.

##### Multiple comparison adjustment

We adopt the randomization-based procedure outlined by Lee et al. ^50^ and implemented in ^33^ to adjust our Fisher-exact p-values for multiple testing, using the following steps:

1. Construct a null randomization distribution with 100, 000 treatment allocations for each of the five cities.
2. Treat each of the 100, 000 test statistics as an “observed test statistic.” Compute unadjusted randomization-based p-values by calculating the proportion of test statistics as or more extreme than the “observed test statistic.”
3. The previous step results in five sets of 100, 000 unadjusted randomization-based p-values from which we retrieve the smallest p-value across all of the cities for each of the 100, 000 iterations.
4. Next, construct a distribution from these minimum p-values. This process captures the joint distribution of the test statistics and rescales hypothetical test statistics to hypothetical p-values of the same 0 − 1 scale. For each city, calculate the adjusted p-value, i.e., the proportion of p-values in this final distribution that are smaller than its unadjusted Fisher-exact p-value.

According to ^50^, randomization-based adjusted p-values account for possible correlations among test statistics and can be more powerful than adjusted Bonferroni p-values. Just as when calculating Fisher-exact p-values, this procedure relies on the assignment mechanism from the original experiment ^51^, and does not require assumptions about the underlying distribution of the data.

#### 2.3.3 Bayesian inference

We use an alternative approach that formally treats the estimation of causal effects as a missing data problem ^52^ and follow the general approach outlined in ^53^. This method imputes the missing vector of potential outcomes, i.e., 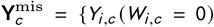 if 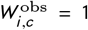 and *Y*_*i,c*_ (*W*_*i,c*_ = 1) if 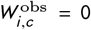}. The Bayesian perspective considers the observed values 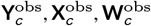 to be a realization of random variables and 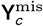 to be unobserved random variables. With this approach, the distribution of the missing potential outcomes can be evaluated by computing the posterior 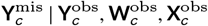. For our purpose of modeling the number of deaths, we propose the following independent count potential outcomes

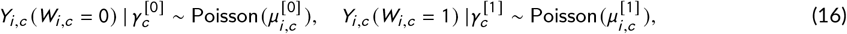

where Poisson denotes the Poisson distribution, and

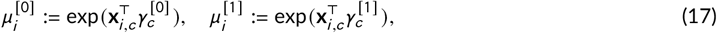

where the covariates **x**_*i,c*_ = (1, *x*_*i*,1,*c*_, …, *x*_*i,H, c*_) are the (*H* + 1)-dimensional vector for each day *i* for city *c* and let 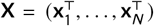 represents the covariates that go into the model, such as temperature and humidity. We also have 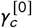 and 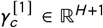. Concretely, we have

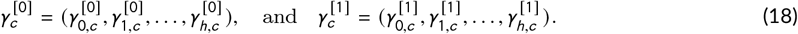

We further assume a prior distribution for ***γ***_*c*_ :

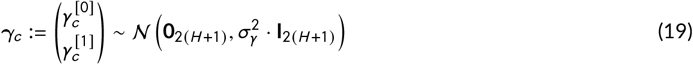

with 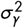 being a fixed positive number, *N* denoting the normal distribution, and ***γ***_*c*_ ∈ ℝ^2(*H* +1)^. Drawing Bayesian inferences for causal effects requires the specification of an assignment mechanism, that is, a probabilistic model for how experimental units are allocated to the treatment assignment given the potential outcomes and covariates. Let **W**_*c*_ denote the *N*_*c*_−vector of treatment assignments with *i*^th^ element *W*_*i,c*_. As before, 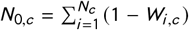 and 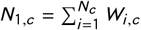 as the number of units assigned to the control and active treatment respectively, with *N*_0,*c*_ +*N*_1,*c*_ = *N*_*c*_. We also assume a paired-randomized assignment design (see Section 4.6, pages 52-53 in ^41^). Using the following identity

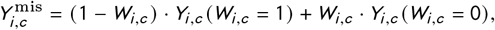

the conditional distribution of 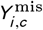 given 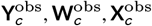 can be expressed as:

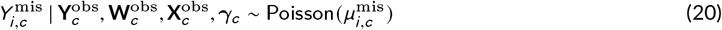

where 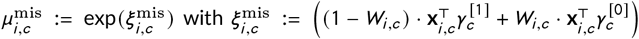. We immediately see from (20) that the conditional distribution of 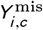 given 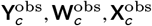 is simply equal to the marginal distribution 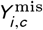 given 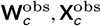 *γ*_*c*_ due to the independence of the potential outcomes. We now proceed to compute the conditional distribution of Y^obs^ given 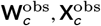, *γ*_*c*_. It follows that

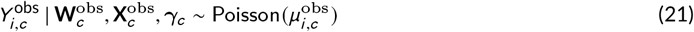

where 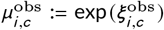 with 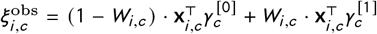. We estimate the average treatment effect (ATE) defined in Equation (3) by repeatedly filling in the missing potential outcomes, which factors in the uncertainty in the imputation. Let 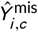 be the imputed value corresponding to 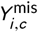, then the ATE in (3) can be estimated as

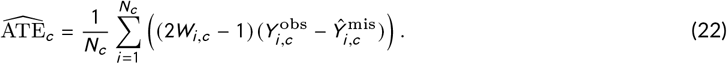

In particular, given a sample of ***γ***_*c*_ (drawn using the simulation methods outlined in Section 8.5^41^), we substitute these values into the conditional distribution of 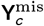 to independently impute all the missing potential outcomes, that is, draw 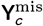 from a Poisson distribution by substituting ***γ***_*c*_ using Equation (20). Substituting the observed and imputed missing potential outcomes into (22) yields an estimate for ATE based on the *r*^*th*^ imputation, 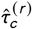 for city *c*:

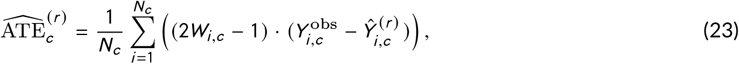

we have used the identity in Equation (20). To derive the full posterior distribution of the ATE, this procedure is repeated *R*_*c*_ times and the average and variance of the imputed estimators 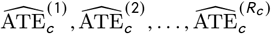 are

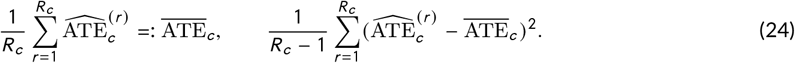

The estimated average treatment effect using the Bayesian approach can be found in Figure 5.

##### Heuristic Connections to Poisson Regression with Interaction Terms

In this section, we compare the traditional Poisson regression framework with a Bayesian approach to estimating the average treatment effect. The methodology closely follows the exposition presented in Chapters 7 and 8 of ^41^, with an important extension: we adapt the analysis to accommodate count outcomes ^53^, rather than the continuous outcomes primarily discussed in their work. From the standard frequentist perspective, the unknown parameters are treated as fixed but unknown quantities, and inference is conditioned upon them. In contrast, the Bayesian framework treats both the missing and observed potential outcomes as random variables—unobserved and observed, respectively—conditional on the model parameters.

Given the specification of the joint distributions *Y*_*i,c*_ (*W*_*i,c*_ = 0) and *Y*_*i,c*_ (*W*_*i,c*_ = 1) in Equation (16), we recall that the average treatment effect is defined as the difference between the two expected values, restated here:

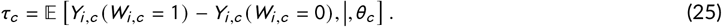

By expanding the terms and leveraging the binary nature of *W*_*i,c*_, we can re-express *µ*_obs_ from Equation (21) as follows:

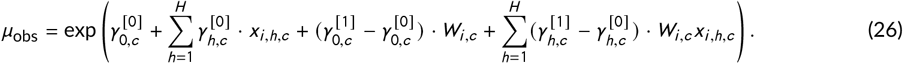

From this, we observe that the average treatment effect (ATE) in Equation (25) can be expressed as:

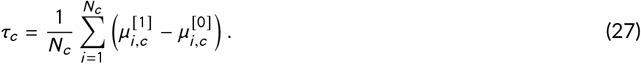

Now let us consider the regression function specified as 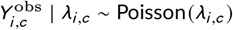, with *λ*_*i,c*_ taking the form:

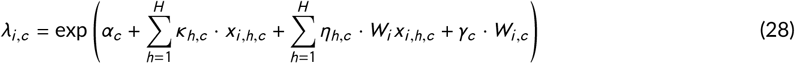

where *α*_*c*_, *γ*_*c*_, *κ*_*h,c*_, and *η*_*h,c*_ denote the regression parameters for city *c*. This specification resembles Equation (2) in Section 2.3.1, but the mean structure corresponds to that of a Poisson regression model. By comparing Equations (26) and (28), we observe that the inferential problem reduces to estimating the regression coefficients via maximum likelihood estimation for the parameters *α*_*c*_, *γ*_*c*_, *κ*_*h,c*_, and *η*_*h,c*_. Once these estimates are obtained, simple transformations can be applied to derive the average treatment effect. Moreover, we note that maximum likelihood estimation under the frequentist framework yields two sets of equations: one corresponding to the estimated control parameters, and another capturing the estimated differences between the treatment and control parameters.

In particular, we have

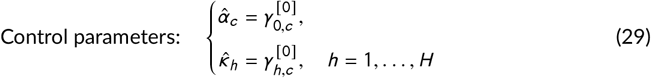

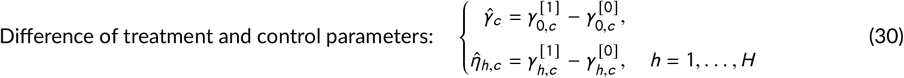

With these, 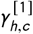 and 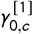 can be constructed from the control parameters and their corresponding treatment-control differences via 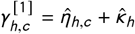 and 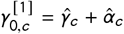, respectively. Equipped with these inferred parameters, we can then compute the ATE in Equation (27) as a function of *µ*^[1]^*i, c* and *µ*^[0]^*i, c* through equations (17) and (18).

### 2.4 Illustrative Example with *H* = 3 Covariates

To concretize the derivations in Section 2.3.1, we present a toy example where the number of covariates is set to *H* = 3. This example demonstrates how the treatment group parameters can be recovered from estimated control parameters and their corresponding treatment-control differences. We then compute the average treatment effect (ATE) accordingly.

#### Example Data

Consider two days with the following covariate values in Table 1.

#### Estimated Parameters

Assume that the following coefficients have been estimated:

- Control group parameters:

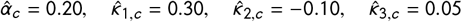
- Differences between treatment and control:

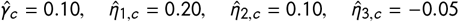

The treatment group parameters are then constructed as follows:

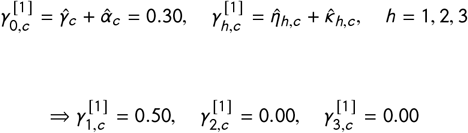

#### Expected Outcomes

Using Equations (17) and (18), we compute the expected outcomes under control and treatment:

- Individual 1:

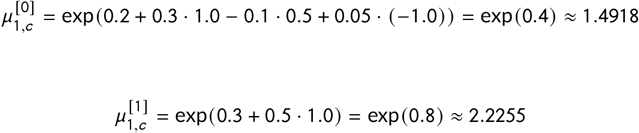
- Individual 2:

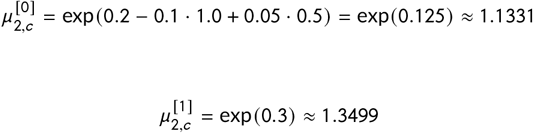

#### Average Treatment Effect

Applying Equation (27), the ATE is given by:

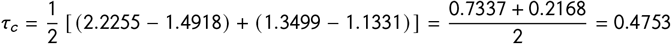

Thus, the estimated average treatment effect in this illustrative case is *τ*_*c*_ ≈ 0.4753.

We defer the theoretical computation of the asymptotic precision of our Poisson regression estimators, as well as the exploration of constraints on precision gains from additional covariates, to future work, following the approaches in ^54^ and ^55^. In the recent paper by ^56^, a causal interpretation of the hazard ratio in randomized clinical trials is provided. Future research may explore connections between the “time series” version of our formulation and theirs.

### 2.5 Stage 4: Interpretation of results

An objective of this causal inference framework is to reduce confounding and draw conclusions not just about the association between heat and mortality, but to be able to assess this causal relationship. We assume that our hypothetical randomized experiment is plausible and that we have accounted for all confounding by matching only on observed covariates ^37^.

It is also important to note that matched treated and control time periods have similar preceding days, but we do not require the three-day treatment periods themselves to be similar. Warm temperatures lead to higher concentrations of many potentially dangerous air pollutants including ground-level ozone, making secondary air pollutants such as ozone possible mediators between heat and mortality ^57^. We then interpret the effect of heat on mortality to include any potential mediators between heat and mortality, such as secondary air pollutants. Finally, these results apply only to the time periods that were selected through the matching process and may not extend to all days in the study period.

### 2.6 Comparison to a traditional analysis of unmatched data

We also compared the results from this causal inference framework to a more traditional analysis; for each city, we fit a Poisson regression model on the full, unmatched dataset. These models have the same form as those in Model 3 (Equation 12) and serve as a point of comparison for the models fit on the matched dataset. We note that these models typically allow for a more complex relationship between exposure to heat and mortality, often with a generalized additive model (GAM) or spline ^58,59^. We use the binary exposure defined in Equation 1 for comparison across models.

## 3 RESULTS

### 3.1 Balance assessment

Here, we present the results of the matching process with covariate balance diagnostics. Note that since we have designed a hypothetical randomized experiment, we seek to identify pairs of treated and control periods with similar lag days, but we do not require days within the three-day experiment to be similar. We seek to balance only the lagged covariates.

First, we must ensure that the matching process leaves us with a reasonable number of pairs in each city for the analysis stage. Table 4 displays the number of 1) control (warm) and treated (hot) units identified, 2) the number of matched pairs of control and treated units, and 3) the percentage of treated units for which a corresponding control unit was available. For the majority of treated units identified in the full, unmatched data, no suitable control units are available for matching within the constraints set in Table 5.

Next, we investigate covariate balance; the goal is that all covariates are balanced in the treated and control groups after matching to minimize potential confounding. Figure 2 is a Love plot, which displays potential relationships between background covariates and the treatment assignment ^60^. For both the full, unmatched data and the matched subset, we calculate the standardized difference in covariate means as follows for each covariate *X*_*i*_.

**FIGURE 2.**
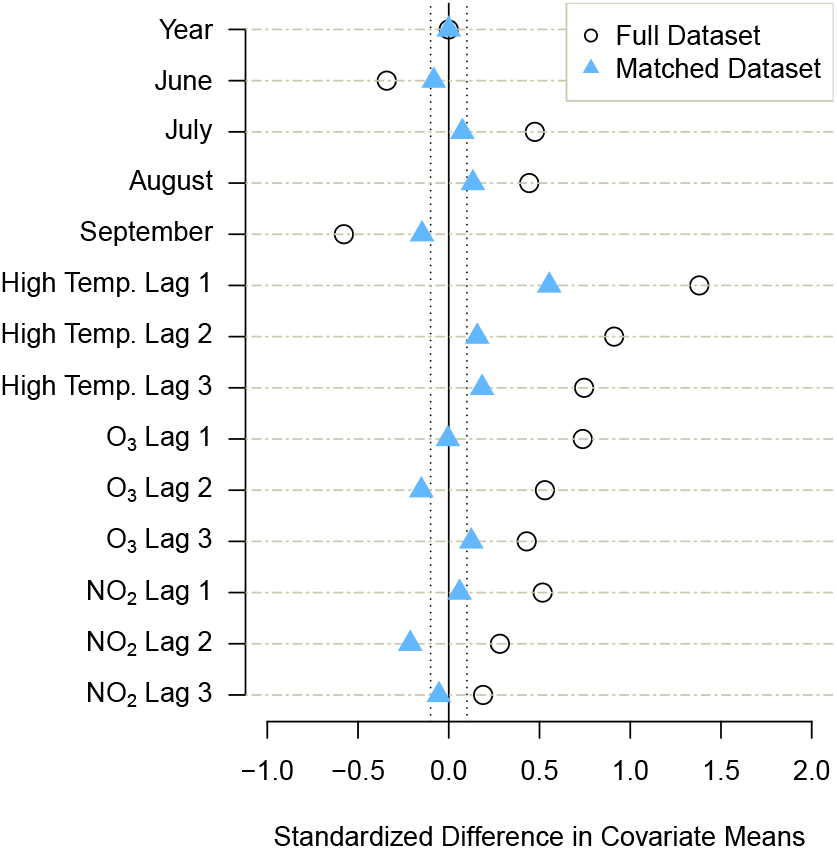
Love plot with the standardized differences in covariate means for several potential confounders; see Equation 31 for this calculation. Standardized mean difference values near zero indicate covariate balance between treated and control units, whereas values that are farther from zero indicate that covariates are associated with treatment assignment. Through matching, we aim to reduce these standardized mean differences to be between −0.1 and 0.1 ^61^, marked here with vertical dashed lines.

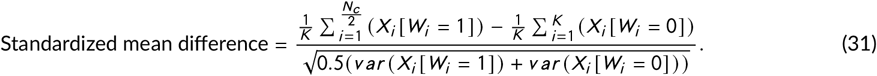

If a covariate is perfectly balanced between treated and control periods, then the covariate has no association with treatment assignment, and the standardized mean difference is 0. We see this with the variable year, which is matched exactly in each pair. A positive standardized mean difference indicates that a covariate is positively associated with assignment to treatment, and a negative standardized mean difference indicates that it is positively associated with assignment to control. We see in Figure 2 that the standardized differences in covariate means were greatly reduced after matching.

We can also inspect density plots to assess covariate balance in continuous covariates that may exhibit complex associations with the treatment assignment. We see in Figure 1 (appendix) that in the full, unmatched data, periods in the middle of the summer were most likely to be hot, while periods in the beginning and end of the summer were most likely to be warm, as expected. However, in the matched dataset, the distribution of hot and warm paired units is more similar throughout the summer, reducing potential confounding by variables with seasonal variations.

We can also visualize these hypothetical randomized experiments by plotting several potential confounders leading up to and during each three-day experimental unit. Figure 3 displays the mean values of six covariates over each three-day treated or control experimental unit, as well as the three days preceding these experimental units. All six covariates display similar patterns. Lag days preceding treated periods have similar covariate values to those preceding control periods. These covariate values then diverge during the three-day experimental units when they are unconstrained by the matching process, thus imitating randomization of treatment assignment on Day 1.

**FIGURE 3.**
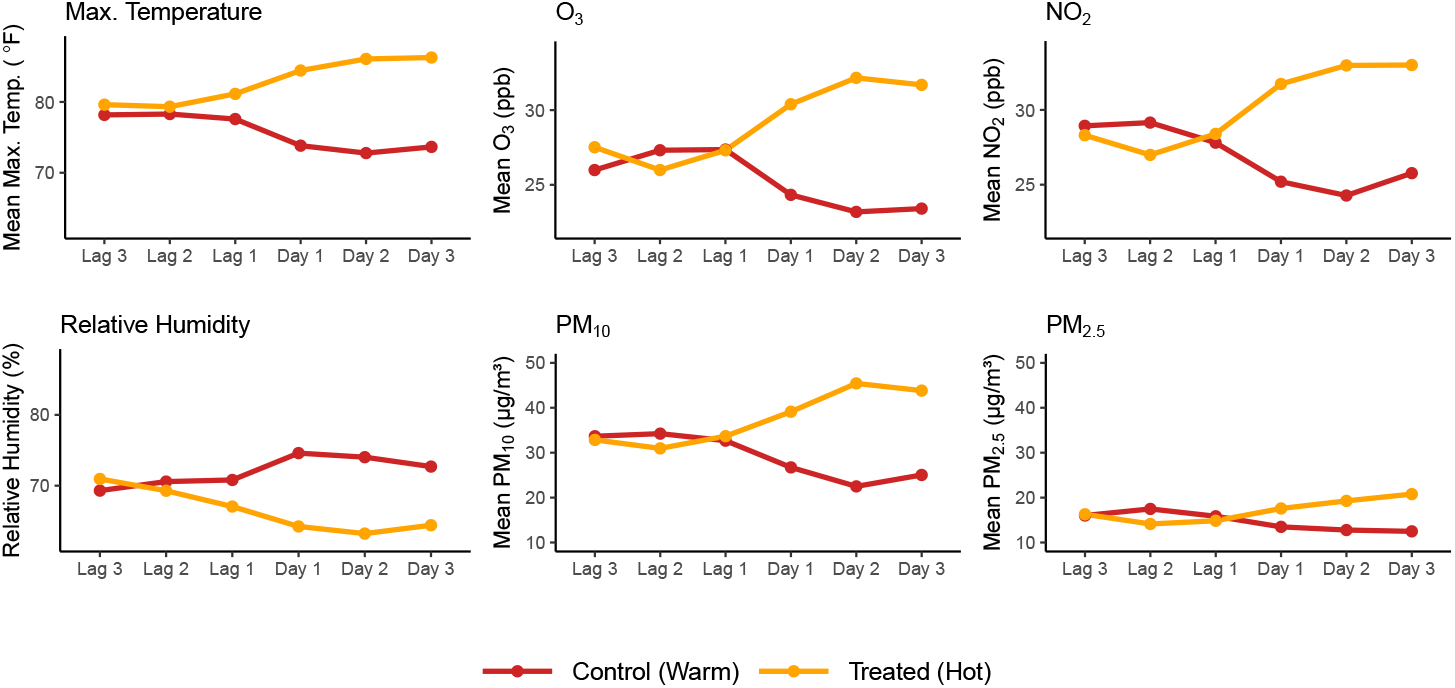
This figure displays the mean values of six covariates in matched treated and control periods over time across all five cities. The first three points in each time series correspond to lag days, or the three days preceding each three-day experimental unit. The last three points in each time series correspond to the three-day treated and control periods in the matched dataset. Note that *PM*_2.5_ and *P M*_10_ were not recorded daily during this time period, so this figure includes only matched periods for which particulate matter concentrations were available. Missingness is assumed to be independent of the exposure assignment.

### 3.2 Three-day mortality during warm versus hot periods

After assessing covariate balance in the matched dataset, we compare mortality during hot versus warm time periods. Figure 4 displays box plots comparing the distribution of three-day mortality counts during matched control and treated periods. In the matched dataset, the median number of deaths during treated periods is greater than the median number of deaths during control periods in all five cities.

**FIGURE 4.**
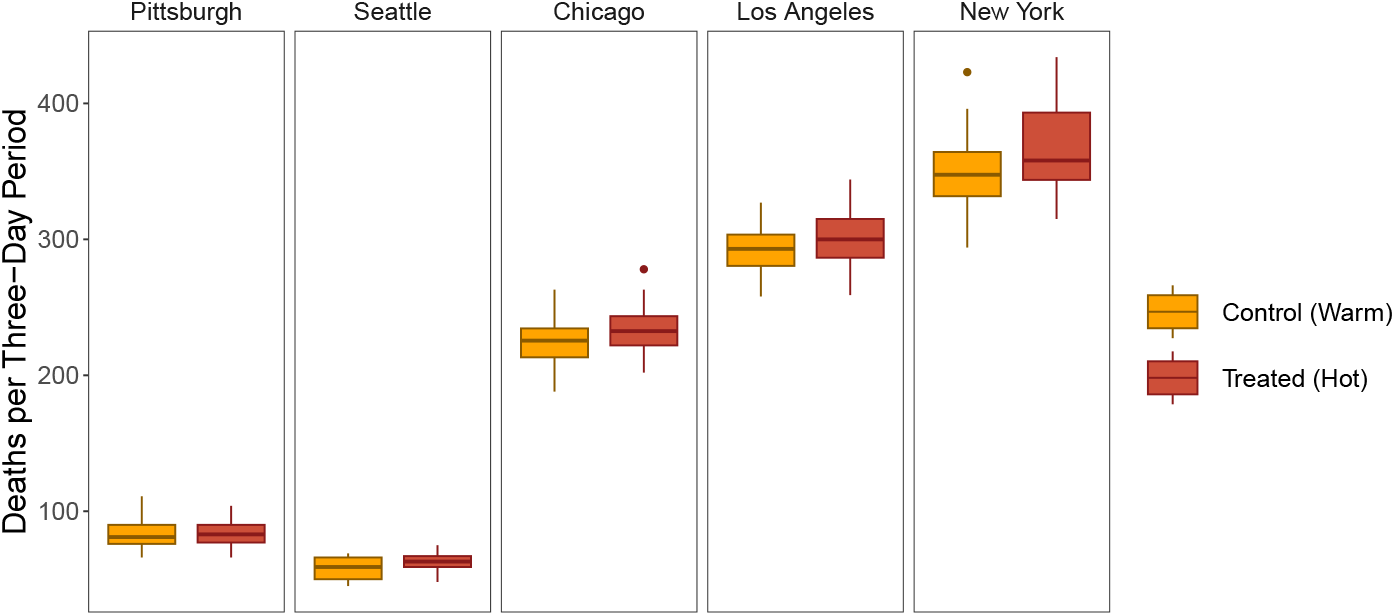
Box plots comparing the distribution of three-day mortality (the total number of deaths over a three-day time period) during matched treated versus control periods.

We display inference results on two scales: 1) the additive scale, as a measure of the difference between the number of deaths during three-day treated and control periods, and 2) the multiplicative scale, as a measure of the ratio between the number of deaths during three-day treated and control periods.

The Neymanian and Bayesian inference approaches described here estimate the average treatment effect *τ*_*c*_ and uncertainty intervals on the additive scale, whereas the traditional frequentist approach of Poisson regression estimates the risk ratio associated with treatment. We also estimated uncertainty intervals for both the estimated average treatment effects on the additive scale and on the multiplicative scale with the Fisherian approach. All inference results on the additive scale are displayed in Figure 5; inference results on the multiplicative scale are displayed in Figure 6.

**FIGURE 5.**
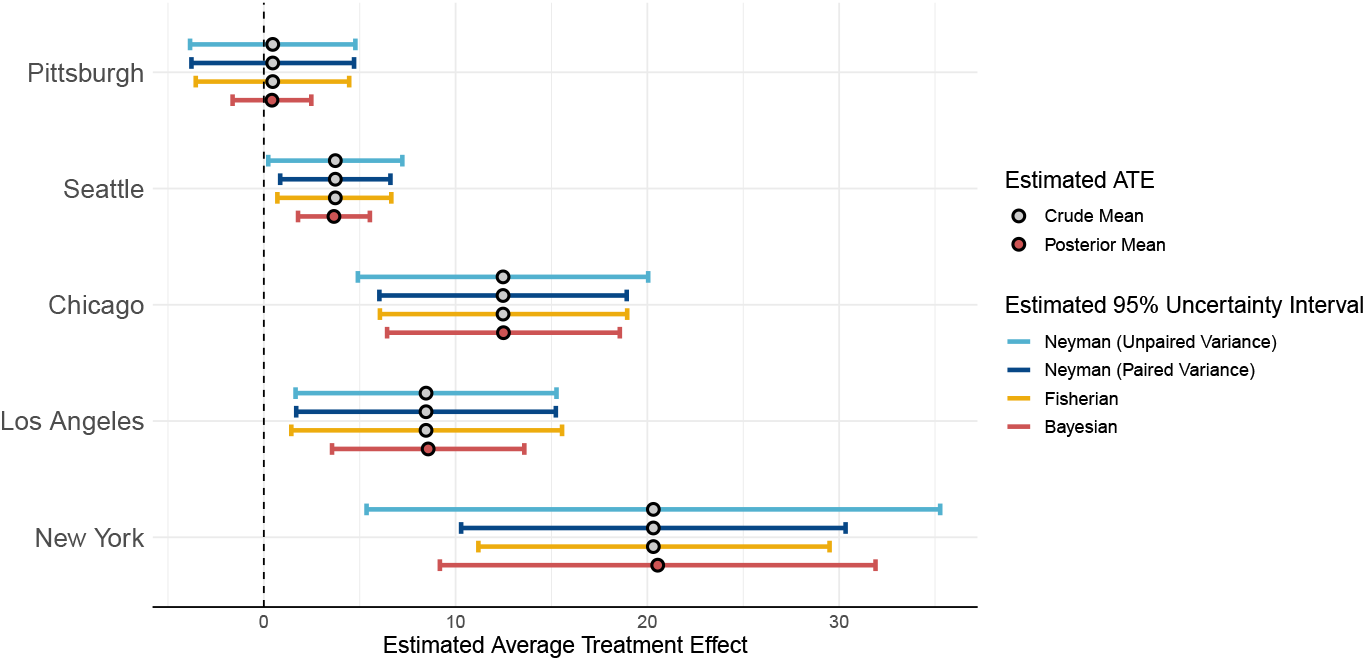
Estimated average treatment effects and estimated 95% uncertainty intervals comparing the total number of deaths during three-day treated and control periods. The Neymanian estimate of the average treatment effect is simply the observed crude mean difference between three-day mortality during treated and control units (Equation 4), while the Bayesian estimate is the posterior mean. See Table 3 (appendix) for exact numerical values displayed here.

**FIGURE 6.**
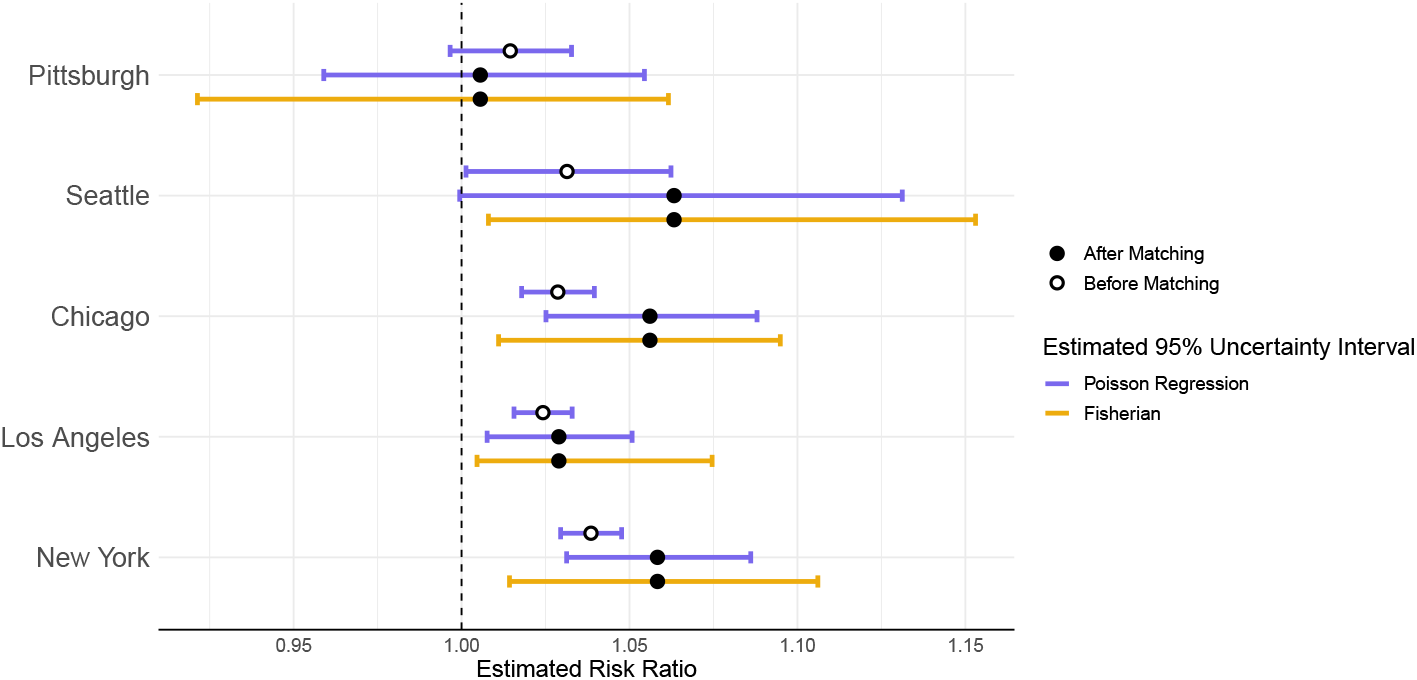
Estimated risk ratios and estimated 95% uncertainty intervals comparing the total number of deaths during three-day treated and control periods. The point estimates represented by open circles were obtained using all three-day periods in the full, unmatched dataset with defined treatment, before matching treated and control periods and eliminating those without corresponding matches, adjusting for potential observed confounders (Model 3). The point estimates represented by closed circles were obtained using the subset of matched treated and control units. Although these point estimates were obtained for Models 1, 2, and 3, only results from Model 1 (the unadjusted model) are displayed here. Fisherian intervals were also obtained for these estimated risk ratios from the matched data. See Table 6 for all exact numerical values displayed in this figure and the results from Models 2 and 3, which were fit on the matched data and were adjusted for certain covariates.

Because the additive scale is dependent on city-specific population, we have included population-adjusted estimates of average treatment effect for each city in Table 3. This table also includes Fisherian p-values for the estimates in each city, as defined in Equation 14.

After matching, we observe more deaths during hot time periods compared to warm time periods among the elderly living in Seattle (3.7, [95%FI: .7, 6.6]), Chicago (12.5, [95%FI: 6.1, 18.9]), Los Angeles (8.5, [95%FI: 1.4, 15.6]), and New York (20.3, [95%FI: 11.2, 29.5]), with unadjusted Fisherian p-values ≈ 0.0078, 0.00005, 0.00848, and < 0.00001, respectively. We retrieve similar results from the Neymanian and Bayesian analyses, which suggests that hotter summer days are associated with higher mortality than cooler summer days in these cities. In Pittsburgh, we could not reject the sharp null hypothesis (Fisherian p-value ≈ 0.42).

The risk ratios obtained from the Poisson regression models align with the results from the other approaches. We found the largest risk ratio in Seattle: 1.076[95%CI: 0.999, 1.16], indicating a 7.6% increase in mortality rates for hot versus warm time periods. This was followed by New York, Chicago and Los Angeles: 1.057[95%CI: 1.023, 1.092], 1.051[95%CI: 1.019, 1.084], 1.039[95%CI: 1.013, 1.065] respectively. For the cities in which we found increases in the deaths (i.e., Seattle, Chicago, New York, and Los Angeles), the risk ratios were larger when using the Poisson regression model on the historical times series matched data compared to those from the Poisson regression model on the unmatched data (i.e., traditional analysis). This finding indicates that the traditional analysis may underestimate the effect of extreme temperatures.

## 4 DISCUSSION

### 4.1 Our results

Our findings indicate meaningful increases in mortality can be attributed to heat in four of the five cities analyzed. This is in agreement with existing literature that has reported on increased mortality due to heat-related episodes. Previous studies conducted on the National Morbidity and Mortality Air Pollution Study (NMMAPS) have estimated increases in mortality risks related to heat waves nationally to be 3%[95% Posterior Interval (PI): 2.4%, 3.6%] and 3.74%[95% PI :2.29, 5.22%] ^59, 62^. Similarly, a county-specific analysis conducted in California found approximately 4% increases in non-accidental mortality in Los Angeles per 10°F, while an 18-year study on New York found that heat waves increased mortality by 6.5% [95% CI: 2.0%, 11.2%] ^63, 64^.

### 4.2 Drawing different types of causal inference

The uncertainty intervals obtained from the frequentist, Fisherian, and Bayesian approaches heavily overlap, indicating similar effect sizes across the three modes of inference. These approaches rely on different assumptions, so their alignment is reassuring. Additionally, three different analysts implemented the different modes of inference separately, highlighting the robust nature of this causal inference analysis and increasing confidence in our results.

Together, the three approaches provide slightly distinct interpretations of the results. Neymanian inference produces an estimated average treatment effect, which we interpret as the estimated average additional number of deaths during a three-day time period had it been hot instead of warm. For example, we observe an estimated average treatment effect of 12.5 deaths in Chicago, indicating that on average, a hot three-day time period will have 12.5 [95%CI: 6, 18.9] more deaths than a warm three-day time period, holding all other observed covariates constant. Similar to Neymanian Inference, the Bayesian models also provide estimates of the average treatment effect. In Chicago, this is estimated to be an increase of 12.495 [95%CI: 6.427, 18.564] more deaths. The intervals obtained from the Fisherian approach have a slightly different interpretation in that they estimate the uncertainty in the unit-level treatment effect consistent with the data as opposed to the average treatment effect. The Poisson regression models provide us with risk ratios; for example, a risk ratio of 1.029 from Model 1 for Los Angeles indicates 2.9%[95%CI: 1.008, 1.051] more deaths during three-day treated time periods compared to control time periods, holding all other observed co-variates constant.

The Neymanian approach assumes a constant additive treatment effect, i.e., the effect of heat is the same across all matched treated and control periods. In order to calculate 95% confidence intervals (CIs), we also assume that the estimated average treatment effect is normally distributed. The Fisherian method allows us to draw inferences without making any asymptotic assumptions. This is particularly useful for small sample sizes and when data do not follow a normal distribution. The Bayesian approach, like the Neymanian approach, estimates the average treatment effect. Here, we specify a model for the joint distribution of (*Y*_*i,c*_ (*W*_*i,c*_ = 0), *Y*_*i,c*_ (*W*_*i,c*_ = 1) and rely on a large sample assumption approximation.

The standard frequentist approach of Poisson regression specifies a model and assumes that mortality counts, conditional on all covariates in the model, follow the Poisson distribution. Poisson regression allows us to estimate risk ratios, and requires additional assumptions to estimate an effect size on the additive scale. All approaches presented here assume independence between time periods, although we expect some temporal correlations within each city. The interpretability of each approach is also an important difference. One benefit of presenting results on the additive scale, i.e., as the estimated average treatment effect or unit-level effect, is that the results may be more interpretable than risk ratios since they quantify the actual number of deaths associated with a certain exposure. We are able to convert results on the multiplicative scale to the additive scale, but only with certain assumptions. This makes the Neymanian, Fisherian, and Bayesian approaches potentially more useful in a policy setting than the frequentist Poisson regression approach.

Had we forgone the matching process entirely and only fit adjusted Poisson regression models on the full, unmatched dataset, it appears as though we may have underestimated the effect of heat on mortality in Seattle, Chicago, Los Angeles, and New York–the four cities in which the effect of heat appears to be significant (Figure 5)–pointing to the importance of matching, although we note that the study population after matching is different.

By constructing a matched dataset prior to our statistical analysis, we mimic a randomized experiment and limit confounding bias common in environmental time series studies. Many of these biases arise from the selection methods for the functions that address confounding in these models. Other biases arise due to unbalanced covariates in the treatment and control groups ^65^. This study’s causal inference framework presents a method for addressing these issues by separating the design from the statistical analysis and is not unique to any particular exposure. This framework can easily be extended to additional environmental health applications where observational data is heavily relied on, or to other applications in time series at large.

### 4.3 Limitations

Although historical time series matching successfully improves observed covariate balance between treatment groups, there are limitations to our approach. Matching requires reducing a dataset down to a smaller subset, which may not be representative of the full population. When we define a dichotomous treatment (Equation 1) and set allowable discrepancies for matching criteria (Table 5), we must ensure that this remaining subset is large enough to draw meaningful conclusions. It is important to note that these results apply only to this subset of matched data in these five cities. Appendix Figure 1 displays covariate differences between the full, unmatched dataset and the matched subset analyzed here.

We also used each city’s summer median maximum temperature to define our dichotomous treatment assignment; we defined a hot time period to be three consecutive days for which the maximum temperature was above the city’s summer median. If we were interested in studying more extreme heat–for example, daily temperature above 90°F, we would require cities experiencing such high temperatures. Additionally, using reported city-wide temperature measurements does not account for neighborhood-level variations in temperature or allow us to evaluate specific impacts on microclimates.

Although previous studies have identified associations between subgroups of the population and increased risk of heat-related mortality ^66^, our data set did not have individual-level (e.g., demographic) information. This information could be particularly helpful for informing policies to mitigate heat-related mortality in susceptible, low-income communities (e.g., programs to expand access to solar-powered air conditioning units in these communities). Additionally, we do not differentiate between mortality caused directly and indirectly by heat. Short-term mortality displacement can also occur when extreme heat expedites time of death in individuals with pre-existing conditions and short life expectancies. This can be seen in a forward shift in the mortality rate followed by a reduction of deaths in the weeks after an extreme heat event. Current literature reports discordant results on the extent to which mortality displacement contributes to heat-induced mortality ^67, 68, 69^. Hence, this question presents a future avenue for exploration.

### 4.4 Relevance of Results

Although not explored within our analysis, our results highlight the importance of government policies and programs that prevent the impact of extreme heat events, which are becoming increasingly common with anthropogenic climate change. Potential strategies include the designation of cooling centers during heat events and programs providing transportation to these locations, which can reduce the risk of overheating, particularly for the susceptible population (e.g., elderly) ^70^. Adopting coordinated heat warning systems can ensure that vulnerable populations have information on upcoming heat events, as well as potential risks and actions to protect themselves ^71^. Future studies could adopt our causal inference framework to evaluate the effects of specific interventions, such as studying random assignments of cooling centers in certain locations to provide further insight on the efficacy of these interventions in reducing heat-related mortality.

It is also important to recognize that the impact of heat extremes can be influenced by spatial factors inherent to the design of a city. Microclimates in urban areas often have higher air temperatures compared to surrounding rural areas due to numerous factors, including lack of vegetation, air pollution, and low reflective materials used in infrastructure ^72^. Increasing the amount of green space in urban areas, as well as the usage of high-albedo materials in houses and buildings to reflect solar radiation can be important in reducing temperatures in cities ^73^. Finally, policies that limit greenhouse gas emissions can mitigate rising temperatures, while those that reduce air pollution can limit the exacerbative effects of these pollutants to minimize heat-related mortality ^74^.

## Data Availability

The data analyzed in this study were derived from the National Morbidity, Mortality, and Air Pollution Study(NMMAPS). These data were publicly accessible at the time they were retrieved. The dataset is no longer available via public repositories.

