## Supplemental File for "Estimating excess mortality during heat waves using a causal inference framework and non-randomized time series"

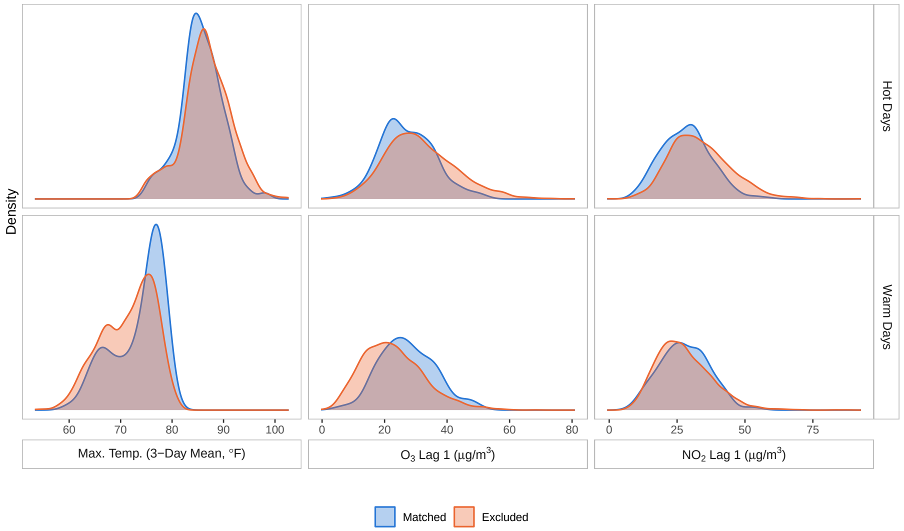


**FIGURE 1** Density distributions of covariates for days included and excluded from the matched sample.

| Day $i$ | $x_{i,1,c}$ | $x_{i,2,c}$ | $x_{i,3,c}$ |
|---|---|---|---|
| 1 | 1.0 | 0.5 | −1.0 |
| 2 | 0.0 | 1.0 | 0.5 |

**TABLE 1** Covariate values for toy example using Bayesian inference.

| City $c$ | **Summer Median Temp$_{max}$** |
| --- | --- |
| Pittsburgh | 80°F |
| Seattle | 72°F |
| Chicago | 80°F |
| Los Angeles | 80°F |
| New York | 82°F |

**TABLE 2** Median daily maximum temperatures measured in each city in June, July, August, and September during the study period (1987-2000).

| | Population 65+ (1990) | Est. ATE | Est. ATE/100k people | Adj. Fisher p-value |
|---|---|---|---|---|
| Pittsburgh | 66,278 | 0.5 | 0.754 | 0.93194 |
| Seattle | 98,605 | 3.7 | 3.752 | 0.03782 |
| Chicago | 403,640 | 12.5 | 3.097 | 0.00022 |
| Los Angeles | 425,219 | 8.5 | 1.999 | 0.04046 |
| New York | 1,186,255 | 20.3 | 1.711 | <0.00001 |

**TABLE 3** This table displays city populations of those 65 years or older in the 1990 census [75], estimated average treatment effects, estimated average treatment effects per 100,000 people, and adjusted Fisher exact p-values.

| | $N_{\text{Control}}$ | $N_{\text{Treated}}$ | $N_{\text{Pairs}}$ | **% Treated Units Matched** |
|---|---|---|---|---|
| Pittsburgh | 500 | 485 | 41 | 8.5% |
| Seattle | 479 | 436 | 33 | 7.6% |
| Chicago | 469 | 467 | 38 | 8.1% |
| Los Angeles | 590 | 472 | 59 | 12.5% |
| New York | 515 | 414 | 32 | 7.7% |

**TABLE 4** This table displays the number of control ($N_{\text{Control}}$) and treated ($N_{\text{Treated}}$) periods identified based on the definitions set in Equation 1, the number of control and treated pairs that were matched by the maximum bipartite matching algorithm ($N_{\text{Pairs}}$), and the percentage of treated units for which an appropriate control unit match was identified ($N_{\text{Pairs}}/N_{\text{Treated}}$).

| Covariate | Discrepancy Allowed |
|---|---|
| Year | 0 |
| Week of the year | ±3 |
| Day of the week | 0 |
| $NO_2$ Lag 1 | ±5 $\mu g/m^3$ |
| $O_3$ Lag 1 | ±5 $\mu g/m^3$ |

**TABLE 5** Maximum discrepancies allowed between a treated period and a control period in order for these periods to be matched. A discrepancy of 0 indicates exact matching.
